# Assessment of Ultra Processed Foods consumption in Senegal: Validation of the Nova-UPF screener

**DOI:** 10.1101/2023.06.26.23291903

**Authors:** Saliou Diombo Kébé, Adama Diouf, Papa Mamadou Dit Doudou Sylla, Kalidou Kane, Caroline dos Santos Costa, Fernanda Helena Marrocos Leite, Giovanna Calixto Andrade, Abdou Badiane, Jean-Claude Moubarac, Nicole Idohou-Dossou, Carlos Augusto Monteiro

## Abstract

**Background:** Ultra-processed foods (UPF), as proposed by the Nova food classification system, are linked to the development of obesity and several non-communicable chronic diseases and deaths from all causes. The Nova-UPF screener developed in Brazil is a simple and quick tool to assess and monitor the consumption of these food products. The aim of this study was to adapt and validate, against the 24-hour dietary recall, this short food-based screener to assess UPF consumption in the Senegalese context.

**Methods:** The tool adaptation was undertaken using DELPHI methodology with national experts and data from a food market survey. The validation study was conducted in the urban area of Dakar in a sample of 301 adults, using as a reference the dietary share of UPF on the day prior to the survey, expressed as a percentage of total energy intake obtained via 24-hour recall. Association between the Nova-UPF score and the dietary share of UPF was evaluated using linear regression models. The Pabak index was used to assess the agreement in participants’ classification according to quintiles of Nova-UPF score and quintiles of the dietary share of UPF.

**Results:** The results show a direct association (p-value<0.001) between intervals of the Nova- UPF score and the average dietary share of UPF. There was a near perfect agreement in the distribution of individuals according to score’s quintiles and UPF dietary share quintiles (Pabak index = 0.84).

**Conclusion:** The study concluded that the score provided by the Nova-UPF screener adapted to the Senegalese context is a valid estimate of UPF consumption.

## Introduction

The Nova system classifies all foods and beverages according to the nature, extent and purpose of the industrial processes they undergo. Food processing includes all physical, biological, and chemical techniques used after food is extracted from production and before it is consumed or made into dishes and meals in kitchens [1]. Foods and beverages are classified according to the Nova system into four groups including Ultra-Processed Foods (UPF). UPF are defined as formulations of ingredients, mostly of exclusive industrial use, that result from a series of industrial processes. Ingredients include derived substances (e.g., emulsifiers, hydrogenated oils) with little or no whole foods to which flavours, colours and other cosmetic additives are added. As a group, UPF have a poor nutritional profile and share characteristics that favour overconsumption such as being hyperpalatable and addictive [2–3]. A meta-analysis involving many representative studies and several other epidemiological studies have shown the negative effect of high UPF consumption on the nutritional quality of diets [4] and the effective link between UPF consumption and the development of obesity and chronic non-communicable diseases (NCDs) such as type II diabetes, hypertension, cardiovascular diseases and all-cause mortality [5–9].

The consumption of UPF is rapidly increasing in low and middle-income countries (LMICs) and this transition, strongly linked to the industrialization, globalization and market deregulation of food systems, represents today a major threat to public health [10–12]. Such as for Senegal, which is facing a rise in the prevalence of overweight and obesity due to lifestyle changes, including sedentary lifestyle and physical inactivity, and unhealthy eating habits. In 2015, the prevalence of overweight and obesity was 22.1% and 6.4%, respectively, among people aged 18 to 69 years. The prevalence of hypertension at national level was 29.8%, and 19.2% and 2.1% of adults had hypercholesterolemia and type II diabetes, respectively in the same period [13].

The Senegalese public health system regularly conducts surveys to assess the state of malnutrition in all its forms, the dietary diversity and micronutrient intakes, but thus far none specifically addresses UPF consumption. Although investigation on UPF consumption and their effects on health in lower-income settings is urgent, researchers and policy makers can find it difficult to monitor consumption trends of UPF. This is mainly because assessing the dietary contribution of UPF using quantitative data, based on 24-hour recall or semi-quantitative food frequency data, is expensive, time consuming and complex to analyse. There is also a considerable lack of data on new dietary patterns and sales and/or consumption trends of ultra-processed foods in these contexts.

The Nova-UPF screener developed and validated in Brazil is a short food-based questionnaire that specifically addresses the consumption of UPF. It asks about UPF consumption (“yes” or “no” question) in the previous day and assigns a score ranging from 0 to 23 according to a list of 23 predefined UPF subcategories. It is a simple and quick tool to administer, requires a low workload, is easy to use and its application will allow the monitoring and evaluation of UPF consumption and possibly their impact on the development of obesity and diet-related NCDs [14–15]. The aim of this study was to describe the adaptation of the Nova-UPF screener to the Senegalese context and evaluate the performance of the score obtained by applying the adapted tool in comparison with the daily energy contribution of UPF obtained from 24-hour dietary recalls as a reference measure.

## 2. Materials and Methods

### 2.1. Ethics

This study received approval (Protocol SEN21/17 - 00000079MSAS/CNERS/SP) from the National Committee of Ethics for Research in Health (CNERS) of the Ministry of Health and Social Action (MSAS). Free and informed consent was obtained from the participants before starting the study.

### 2.2. Adaptation of the Nova-UPF Screener

The original version of the Nova-UPF screener was developed in Brazil and validated against a full 24-hour recall [15]. To be applicable in our context, the original content (Table 1) was adapted and tested to reflect Senegalese dietary patterns by the Human Nutrition and Food Research Laboratory (LARNAH) team and its partners, including researchers who worked on the development of the original version (Center for Epidemiological Research in Nutrition and Health, NUPENS). The changes made to the original version of the Nova-UPF screener were carried out through a collaborative process, led by the LARNAH research team with key stakeholders in the domains of nutrition, epidemiology, public health and food technology at national level. First, a literature review was performed on limited existing data, particularly Euromonitor reports of countries from the same economic zone and with a similar food environment (e.g., Ghana and Côte d’Ivoire) [16–17], to map the availability of UPF and carry out a first content adjustment. Then, two virtual workshops and individual consultation sessions involving the aforementioned stakeholders were organised (further details below). During the workshops, the DELPHI method was applied, which aims to gather expert opinions on a specific subject and to highlight convergences and consensus on the orientations to be given to a project by subjecting these experts to successive waves of questions, was applied [18].

**Table 1:**
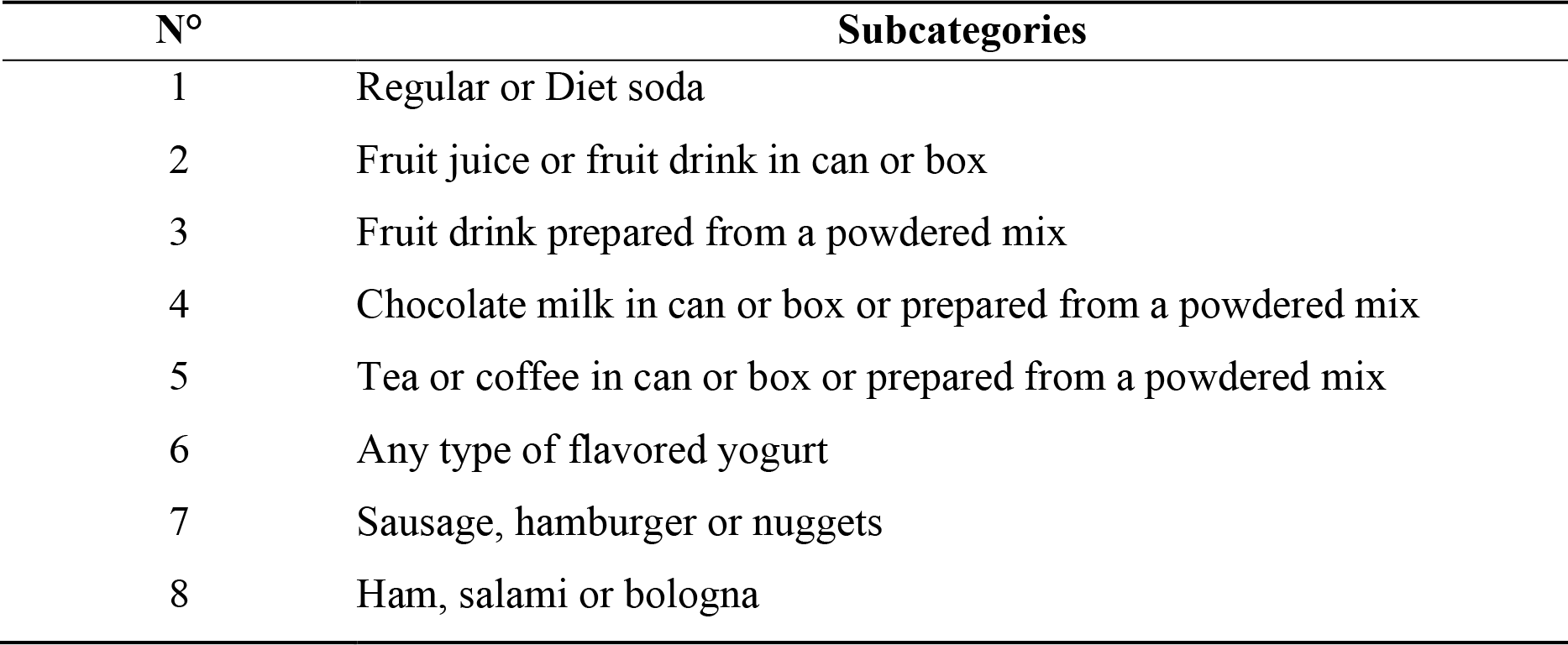
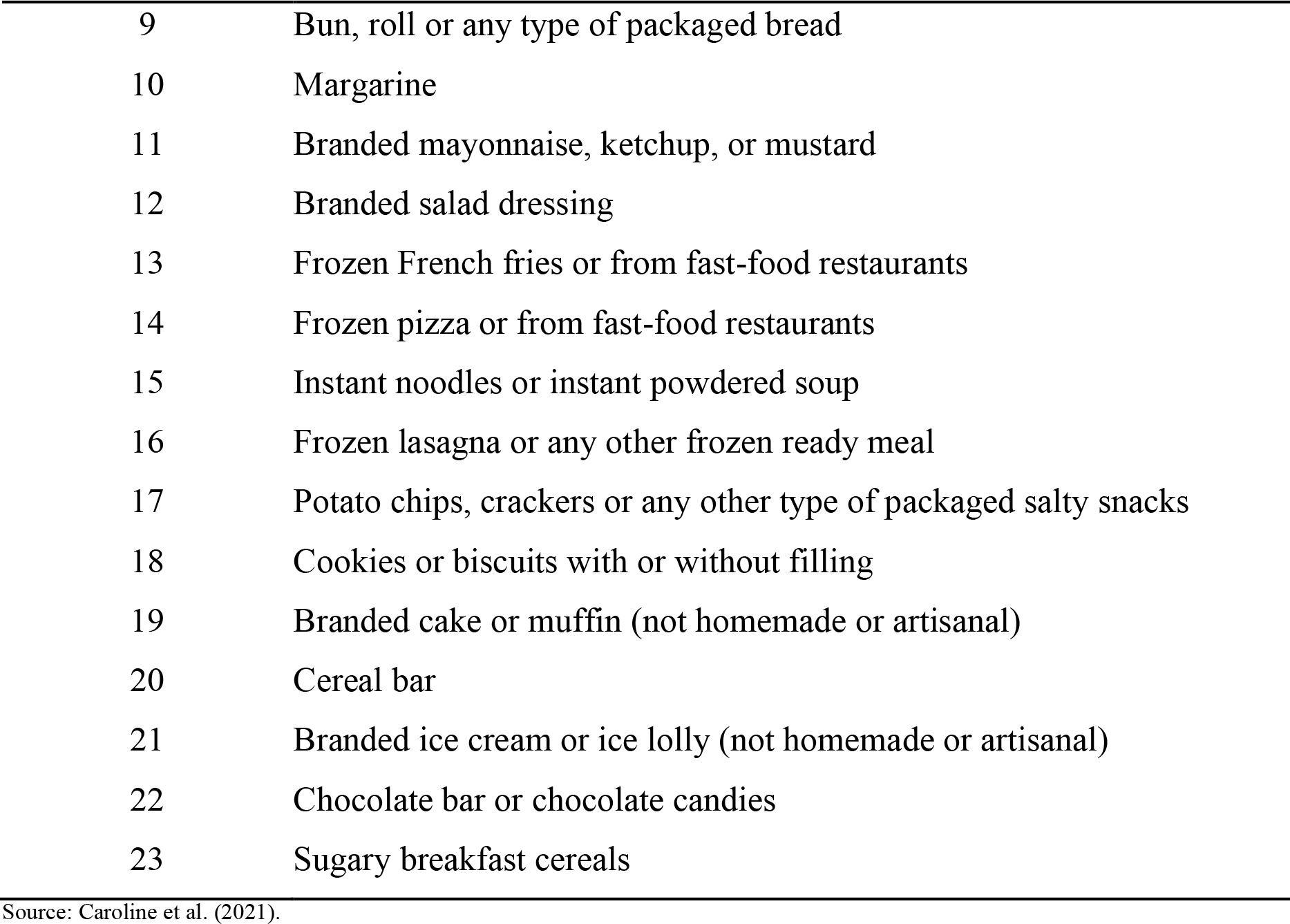
Original version of the Nova-UPF screener.

| N° | Subcategories |
| --- | --- |
| 1 | Regular or Diet soda |
| 2 | Fruit juice or fruit drink in can or box |
| 3 | Fruit drink prepared from a powdered mix |
| 4 | Chocolate milk in can or box or prepared from a powdered mix |
| 5 | Tea or coffee in can or box or prepared from a powdered mix |
| 6 | Any type of flavored yogurt |
| 7 | Sausage, hamburger or nuggets |
| 8 | Ham, salami or bologna |
| 9 | Bun, roll or any type of packaged bread |
| 10 | Margarine |
| 11 | Branded mayonnaise, ketchup, or mustard |
| 12 | Branded salad dressing |
| 13 | Frozen French fries or from fast-food restaurants |
| 14 | Frozen pizza or from fast-food restaurants |
| 15 | Instant noodles or instant powdered soup |
| 16 | Frozen lasagna or any other frozen ready meal |
| 17 | Potato chips, crackers or any other type of packaged salty snacks |
| 18 | Cookies or biscuits with or without filling |
| 19 | Branded cake or muffin (not homemade or artisanal) |
| 20 | Cereal bar |
| 21 | Branded ice cream or ice lolly (not homemade or artisanal) |
| 22 | Chocolate bar or chocolate candies |
| 23 | Sugary breakfast cereals |

Source: Caroline et al. (2021).

The first virtual workshop carried out in June 2021 with over 20 national stakeholders was conducted to i) present the Nova-UPF screener to the audience and ii) define the guidelines for adapting the original content of the Nova-UPF screener to the Senegalese context. These included finding resources to fill the data gap on UPF consumption in the country and to define a methodological approach to revise the proposed subcategories. As a result, we implemented a market survey to characterise the food supply in Dakar (the capital of Senegal) and capture the diversity and main types of ultra-processed foods available in the food environment. The Dakar region concentrates most of the economic activity, flow of food products to the rest of the country, and all major distribution chains [19]. More than 4,700 types of packaged food products, of which more than 70% was ultra-processed, were listed and classified into different subcategories.

Following the first virtual workshop, individual one-hour consultations were held with experts to directly obtain their opinions and suggestions on the content to be adapted (e.g., what types of UPF, which brands or examples should be included). The results of these interviews and data from the market survey were used to develop a first adapted content proposal. The second workshop carried out in September 2021 with the same stakeholders reviewed the suitability of selected UPF subcategories and examples of products included in each subcategory to ensure that they were relevant to the Senegalese context. Following this second consultation with national experts, the final content of the Nova-UPF screener for Senegal was obtained.

Before implementing the tool, face-validity was assessed to evaluate the tool’s design, content, structure, and ease of understanding. The Nova-UPF screener in its original design is a self-administered questionnaire where the respondent reads the names of the subcategories and checks the answers. However, given the low level of literacy and linguistic diversity, the Nova-UPF screener for Senegal was completed with the assistance of a trained interviewer who could explain the tool to respondents, and the meaning of each UPF subcategory, when necessary. In this phase, an electronic questionnaire was designed to allow interviewers to personally assess, after each question (subcategory): i) whether the wording of the requested subcategory was well understood and ii) whether the respondent had no difficulty in answering. Then, participants were asked: i) whether he/she had understood the meaning of each subcategory, ii) whether the proposed UPF subcategories corresponded to the food products to which they were often exposed, iii) whether the examples used were clear and well recognised. Based on these results, further minor adjustments to the phrasing of the questions were made. From the original version, several changes were made. These included rewording of subcategory names, the splitting or grouping of certain subcategories and the introduction of new subcategories. In Figure 1, we have a summary of the changes made from the original version. Like the original tool, the Nova-UPF screener for Senegal also presents 23 subcategories. Foods were grouped within the same subcategory according to their composition or use. At the end of the adaptation phase and content validation (face-validity), a final version of the Nova-UPF screener adapted for Senegal was obtained (see Table 2).

**Figure 1:**
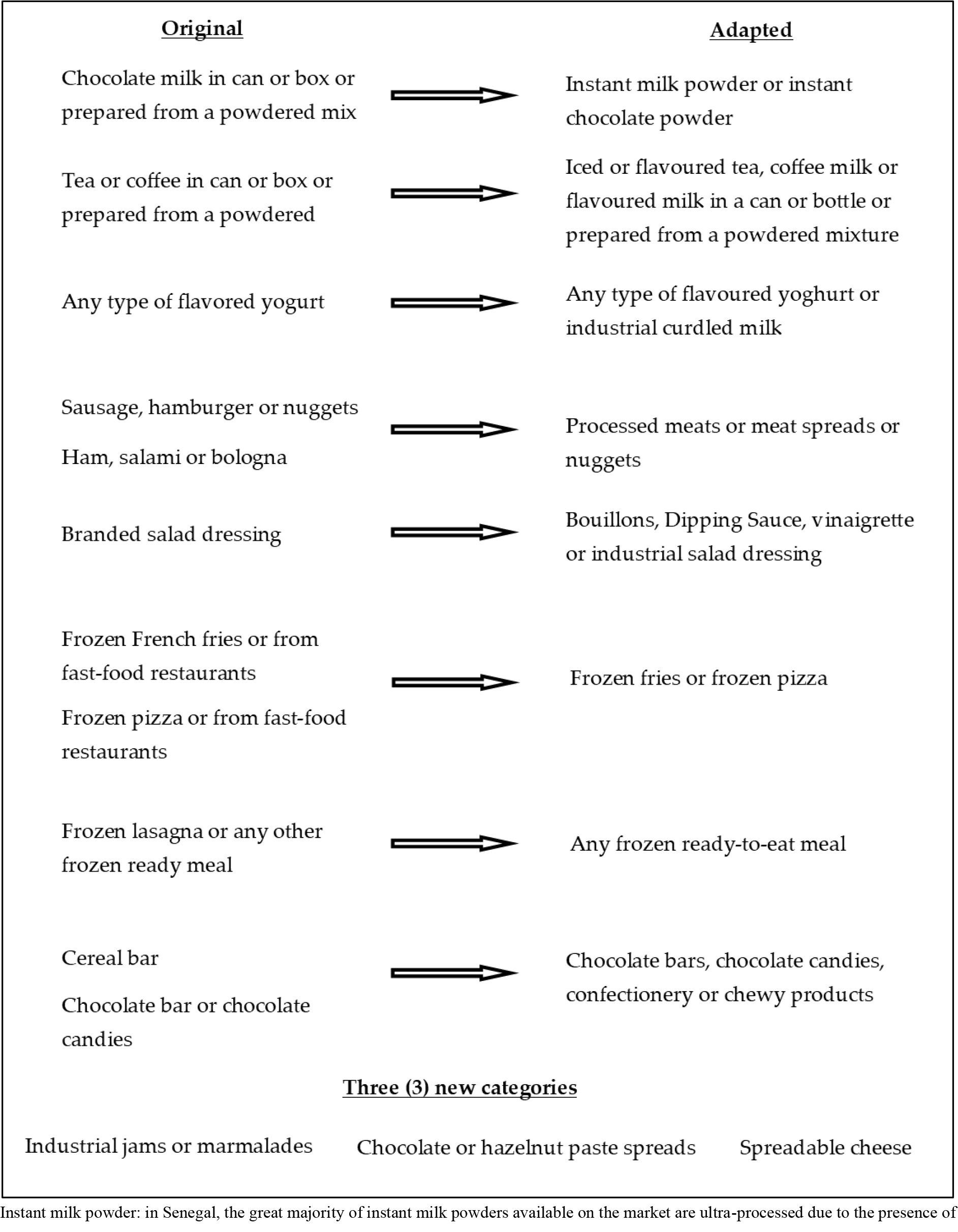
Summary of changes to the original content

**Table 2:** Final adapted version.

| N° | Subcategories | Code |
| --- | --- | --- |
| 1 | Soda, energizing or diet beverages | CAT 01 |
| 2 | Flavoured drinks, concentrates and/or fruit nectars in a can or box | CAT 02 |
| 3 | Fruit-flavoured drinks prepared from a powdered mixture | CAT 03 |
| 4 | Instant milk powder or instant chocolate powder | CAT 04 |
| 5 | Iced or flavoured tea, coffee milk or flavoured milk in a can or bottle or prepared from a powdered mixture | CAT 05 |
| 6 | Any type of flavoured yoghurt or industrial curdled milk | CAT 06 |
| 7 | Processed meats or meat spreads or nuggets | CAT 07 |
| 8 | Bread, rusks, any other type of packaged industrial bread | CAT 08 |
| 9 | Frozen fries or frozen pizza | CAT 09 |
| 10 | Margarine | CAT 10 |
| 11 | Industrial jams or marmalades | CAT 11 |
| 12 | Chocolate or hazelnut paste spreads | CAT 12 |
| 13 | Spreadable cheese | CAT 13 |
| 14 | Industrial mayonnaise, ketchup or mustard | CAT 14 |
| 15 | Bouillons, Dipping Sauce, vinaigrette or industrial salad dressing | CAT 15 |
| 16 | Instant soup powder or instant noodles | CAT 16 |
| 17 | Any frozen ready-to-eat meal | CAT 17 |
| 18 | Chips, crackers, or any other type of packaged salty snack | CAT 18 |
| 19 | Cookies or biscuits with or without fillings | CAT 19 |
| 20 | Industrial cakes, muffins, or pastries | CAT 20 |
| 21 | Sweetened breakfast cereals | CAT 21 |
| 22 | Industrial ice cream or popsicles | CAT 22 |
| 23 | Chocolate bars, chocolate candies, confectionery or chewy products | CAT 23 |

### 2.3. Validation of the Nova-UPF Screener

#### 2.3.1. Design and selection of participants

The validation study involved adults aged 18 years and older, living in urban areas of Dakar, which concentrates nearly a quarter of the national population and almost half of the country’s urban population (49.6%) [20]. For validation studies, higher numbers of subjects will provide better estimates of reproducibility or validity and a sample size of at least 50 to100 subjects is recommended [21].

A two-stage random sampling design was used. First, a random selection was taken in 25 urban census districts (CD), spread across nine (9) health districts in Dakar’s medical region. Second, 12 households were randomly selected in each CD (13 in one of them), by using a sampling step defined according to the size of the CD. Finally, in each household, one adult was randomly selected, taking into account a 50/50 balance between men and women. In total, 301 adults were enrolled to participate in this study.

#### 2.3.2. Data collection

The data collection was carried out on December 2021, by experienced nutritionists and investigators who were trained in the use of the Nova-UPF screener and quantitative 24h recall. All participants were informed about the aim of the study and invited to join after agreeing to participate by signing the consent form. Then sociodemographic and food consumption data (Nova-UPF screener and 24-hour recall) were collected. The electronic questionnaire was deployed in tablets using the ODK application for data collection (socio-demographic and food consumption), where the 23 subcategories were listed along with a descriptive text of the category and illustrative images as shown in Figure 2. Images corresponding to the mentioned brands were selected taking into account the different forms of packaging (bottle, can, brick sachet, box etc.).

**Figure 2:**
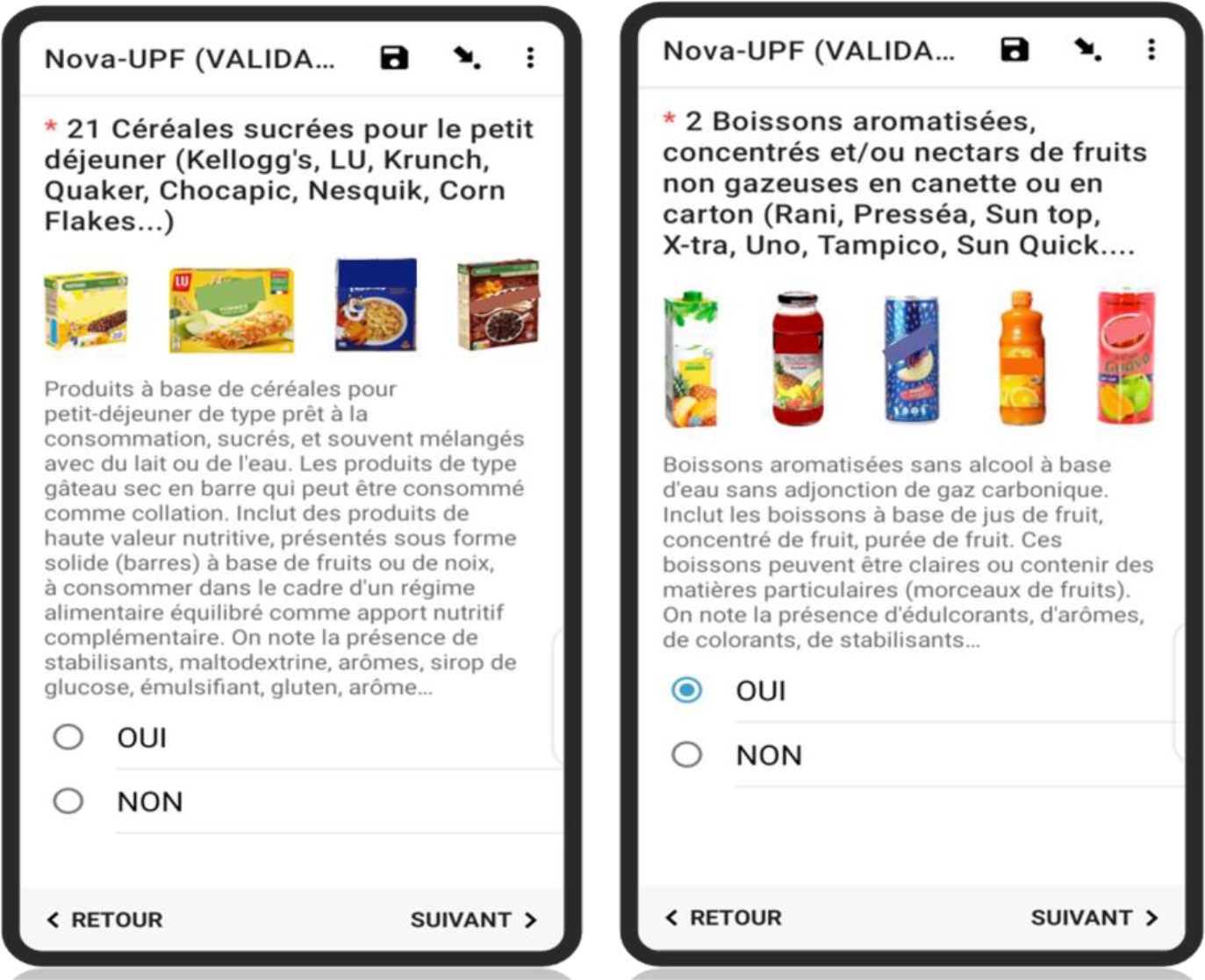
Electronic version of the Nova-UPF screener for Senegal.

The Nova-UPF screener was administered first, for an average time of five minutes. After completion of the tool, investigators conducted the 24-hour dietary recall, where participants informed about all foods and drinks and beverages, and total amount consumed on the day before.

The 24-hour dietary recall was conducted using the automated multiple-pass method [22]. First, participants report, in a rapid and uninterrupted manner, all foods and beverages consumed. Next, the interviewer asks, from an initial list of often forgotten foods, what other foods or beverages the respondent might have omitted to report. The participant is then asked about the type, time and place of each meal, and then provides details such as the method of preparation, quantities and the addition of other foods (e.g., sugar, salt, etc.). Finally, the interviewer lists all the foods previously reported by the respondent, reviewing any omissions. Given local food habits, where people are used to eat the main meals together around the same bowl, the quantity of food or meals consumed was estimated by direct weighing (if a replica was available in the household), by non-standard measurements (e.g., slice, unit, tablespoon, bag, handful, etc.), or by using substitutes (water, dry millet, modelling paste) to get the weight that refer to the volume occupied by the amount of food consumed.

### 2.4. Data analysis

The Nova-UPF score of each participant, which corresponds to the total number of subcategories (from 0 to 23) consumed on the day before, was calculated. To estimate the dietary share of UPF, each food reported in the 24-hour recall was first classified according to the Nova food classification system as 1) unprocessed or minimally processed, 2) processed culinary ingredients, 3) processed foods or 4) ultra-processed foods [1–2]. Then, the real amount consumed of each food was obtained using the database of food conversion factors and the Senegalese standard recipe database, developed under the national food consumption survey implemented by *Consortium pour la Recherche Économique et Sociale* (CRES) in collaboration with LARNAH, the Ministry of Health and the Food and Agriculture Organization of the United Nations in Senegal (FAO-Senegal). These conversion factors are coefficients that allow us to move from non-standard measurements or substitute quantities to real quantities of food. The amount of each food (in grams) was converted to calories using the 2019 West African Food Composition Table [23] and the CIQUAL 2020 Nutritional Food Composition Table [24]. Finally, for each person, we calculated the total calories consumed on the day prior to the interview, including the total calories from UPF and also the percentage of calories from UPF.

To test the association between the score provided by the Nova-UPF screener and the dietary share of UPF obtained from the 24-h recall two methods were used. First, the variation in the average percentage of calories from UPF was examined according to the changes in the score quintiles on a continuous basis and also by comparing the quintiles of distribution of the two variables. In both cases, linear regression models were used to estimate the association. The level of agreement between the quintiles for the score and the percentage of calories from UPF was assessed by calculating the prevalence and bias-adjusted Kappa index (Pabak) [25]. Values greater than 0.80 indicate near-perfect agreement; between 0.61 and 0.80, substantial agreement; between 0.41 and 0.60, moderate; between 0.21 and 0.40, fair; and equal to or less than 0.20, slight [26]. Data analyses were performed with the Stata® 16.1 software and the Pabak index was calculated using R Studio software.

## 3. Results

### 3.1. Socio-demographic characteristics

The Sociodemographic characteristics of participants are described in Table 3. The mean age of subjects was 42 ± 14 years. All 301 volunteers in this study were aged between 20 and 68 years, and the most represented age group was those of 20-39 years. More than half of individuals were married (61.5%) and had reached high school or university (52.8%). Regarding their occupation, 22.9% were in the trade sector, and 11% were unemployed, including 18.7% of women.

**Table 3:** Socio-demographic characteristics (n=301). Senegal (2021).

|  | **Total**<br>**(n= 301)** | **Men**<br>**(n= 151)** | **Women**<br>**(n= 150)** |
| --- | --- | --- | --- |
| **Mean age** | 42 ± 14 | 41 ± 14 | 43 ± 15 |
| **Age group** |  |  |  |
| 20 – 39 | 49.5 (149) | 55 (83) | 44 (66) |
| 40 – 59 | 34.2 (103) | 31.1 (47) | 37.3 (56) |
| 60 – 68 | 16.3 (49) | 13.9 (21) | 18.7 (28) |
| **Literacy** | 77.1 (232) | 82.8 (125) | 71.3 (107) |
| **Level of Education** |  |  |  |
| Primary school | 21.6 (65) | 13.9 (21) | 29.3 (44) |
| College / high school | 24.2 (73) | 26.5 (40) | 22 (33) |
| University | 28.6 (86) | 25.8 (39) | 31.4 (47) |
| Vocational school | 16.3 (49) | 21.2 (32) | 11.3 (17) |
| Others | 9.3 (28) | 12.6 (19) | 6 (9) |
| **Marital status** |  |  |  |
| Single | 23.6 (71) | 31.1 (47) | 16 (24) |
| Divorced | 4.6 (14) | 3.3 (5) | 6 (9) |
| Married monogamous | 51.5 (155) | 55 (83) | 48 (72) |
| Married polygamous | 14 (42) | 10.6 (16) | 17.3 (26) |
| Widow(er) | 6.3 (19) | 0 (0) | 12.7 (19) |
| **Occupation** |  |  |  |
| Unemployed | 11 (33) | 3.3 (5) | 18.7 (28) |
| Student | 9.6 (29) | 8.6 (13) | 10.7 (16) |
| Manual worker | 7 (21) | 13.3 (20) | 0.7 (1) |
| Trader | 22.9 (69) | 13.3 (20) | 32.7 (49) |
| Housekeeper | 11.6 (35) | 2 (3) | 21.3 (32) |

The results are expressed as percentages with the effective (n) or mean ± SD.

### 3.2. UPF consumption and distribution of the Nova-UPF score

As described in Figure 3, the most consumed UPF subcategories were Bouillons, dipping sauce, vinaigrette or industrial salad dressings (32.5%); Industrial mayonnaise, ketchup or mustard (26.3%); Instant milk powder or instant chocolate powder (14.5%); Margarine (6.6%), and Chocolate bars, chocolate candies, confectionery or chewy products (4.9%). Overall, no products belonging to the subcategories Fruit-flavoured drinks prepared from a powdered mixture (CAT 03); Industrial jams or marmalades (CAT 11); and Industrial ice cream or popsicles (CAT 22) were consumed by our study sample.

**Figure 3:**
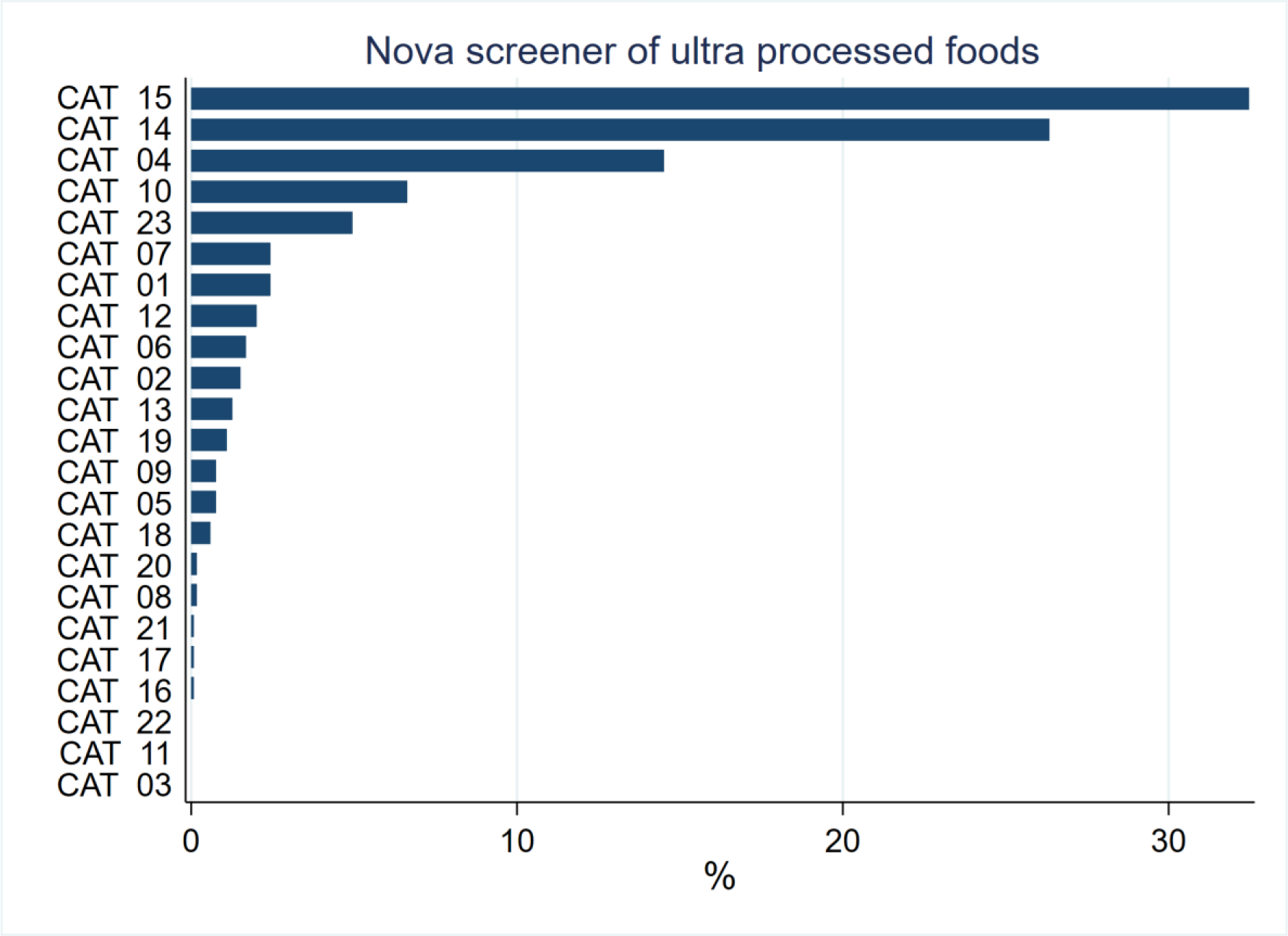
Consumption frequencies of ultra-processed food subcategories.

The Nova scores ranged from zero (0) to eight (8) (Figure 4). The majority of participants got scores of 3 (28.6%), 2 (27.2%), 1 (20.3%), and 4 (13.3%). Only one individual (0.3%) had a score of zero and 10.3% of individuals had a score of 5 or higher (≥ 5).

**Figure 4:**
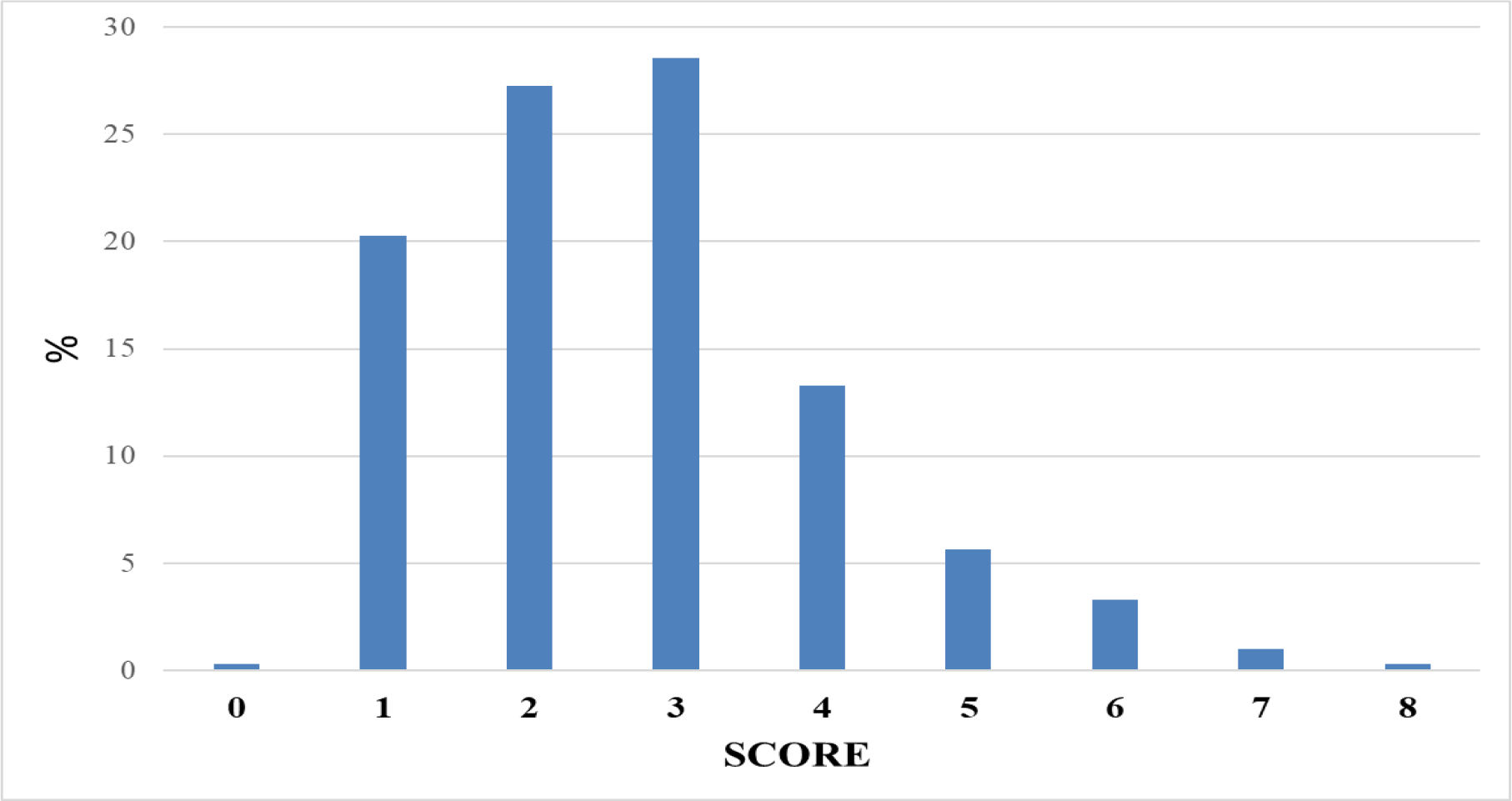
Nova-UPF score distribution.

### 3.3. Dietary share of ultra-processed foods according to Nova score intervals

Overall, the average contribution of UPF to daily energy intake was 17.4% (CI= 18.5 - 21.3). The results showed that the dietary share of UPF consumption increased linearly with the increase in the intervals of the Nova-UPF score (p-value for linear trend <0001) (Figure 5). Individuals in the first quintile had a mean energy contribution from UPF lower than 3%, while the mean energy contribution from UPF for those in the fifth quintile was more than 30%.

**Figure 5:**
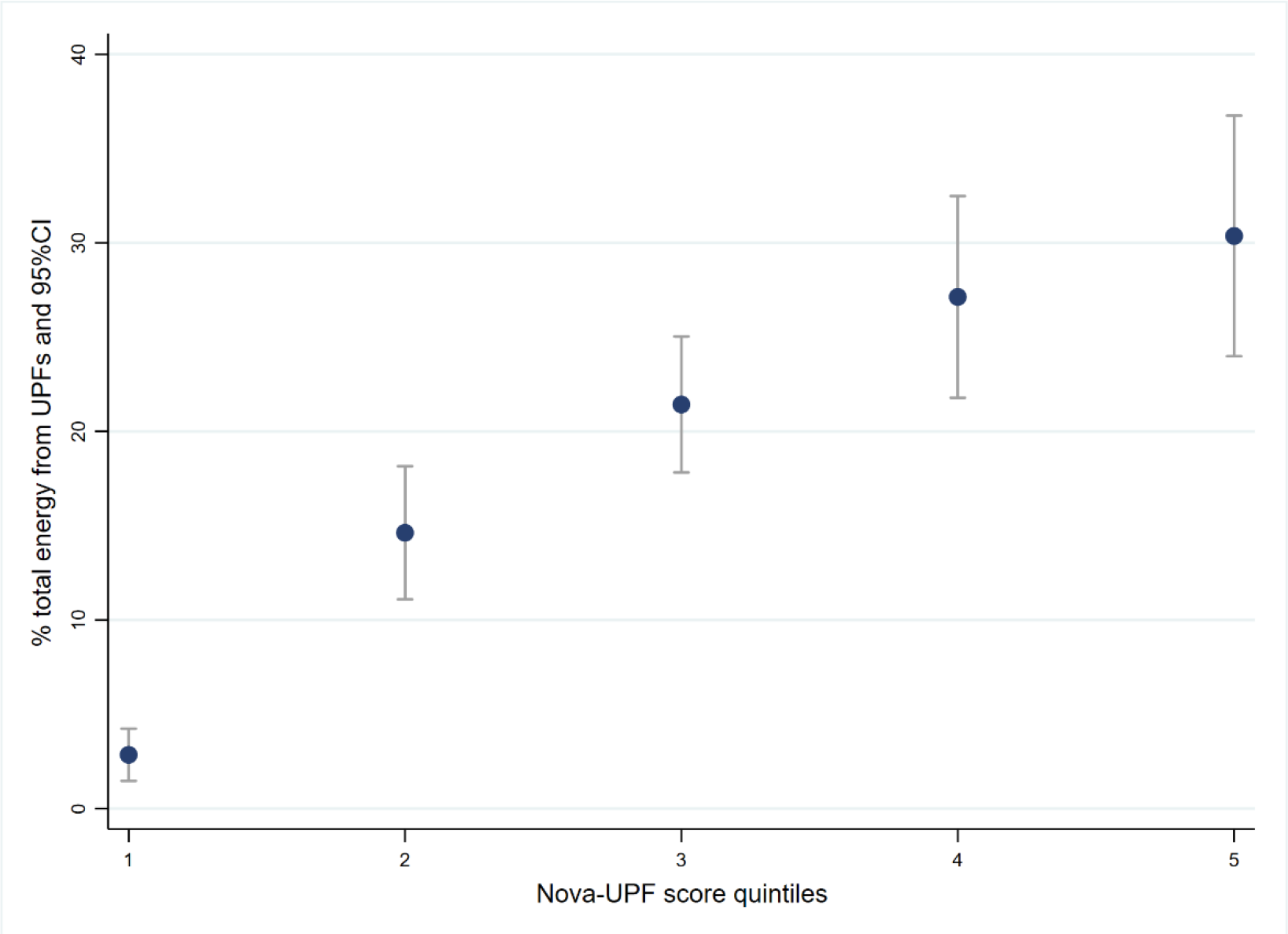
Mean dietary share of UPF obtained from full 24-h recall according to Nova-UPF score intervals

### 3.4. Agreement between the Nova-UPF Screener and the 24-hour dietary recall

Table 4 shows the distribution of individuals according to quintiles of the Nova-UPF score and quintiles of energy contribution from UPF. The Pabak index value was 0.84 (0.68 - 1). This indicates an almost perfect agreement (>0.80) between the two criteria, the Nova-UPF score and the 24-hour dietary recall.

**Table 4:**
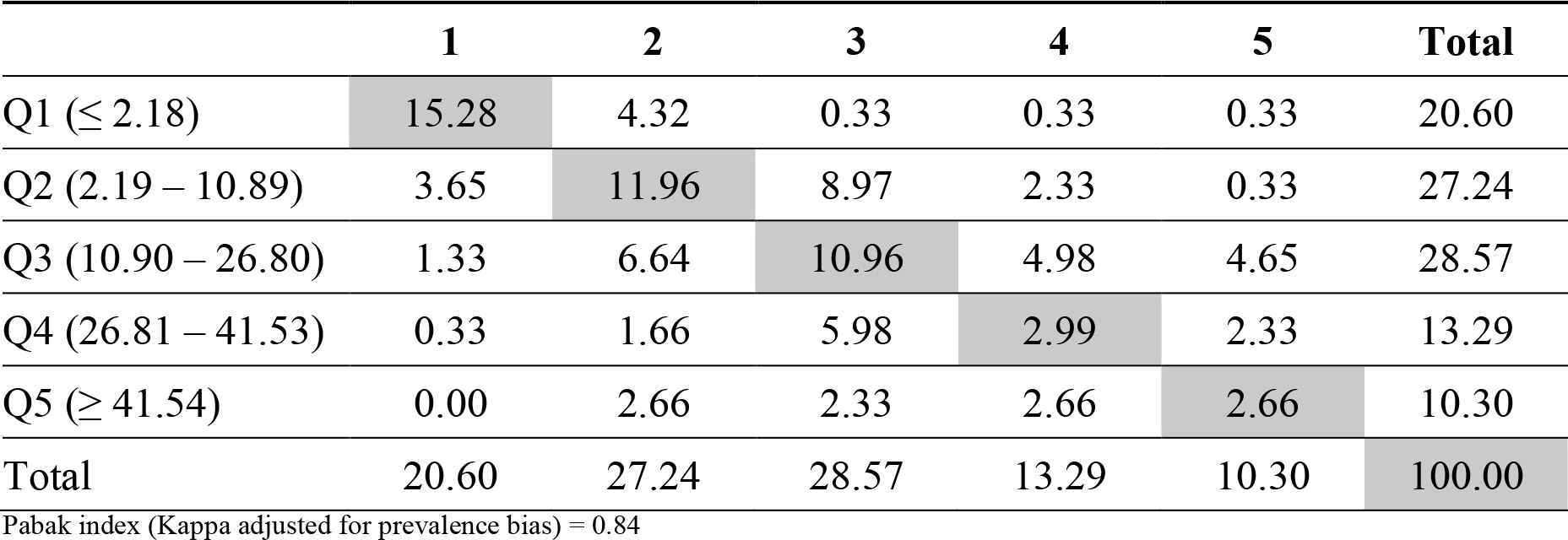
Distribution of individuals by score quintiles and UPF energy contribution quintiles.

|  | 1 | 2 | 3 | 4 | 5 | Total |
| --- | --- | --- | --- | --- | --- | --- |
| Q1 ($\leq 2.18$) | 15.28 | 4.32 | 0.33 | 0.33 | 0.33 | 20.60 |
| Q2 (2.19 – 10.89) | 3.65 | 11.96 | 8.97 | 2.33 | 0.33 | 27.24 |
| Q3 (10.90 – 26.80) | 1.33 | 6.64 | 10.96 | 4.98 | 4.65 | 28.57 |
| Q4 (26.81 – 41.53) | 0.33 | 1.66 | 5.98 | 2.99 | 2.33 | 13.29 |
| Q5 ($\geq 41.54$) | 0.00 | 2.66 | 2.33 | 2.66 | 2.66 | 10.30 |
| Total | 20.60 | 27.24 | 28.57 | 13.29 | 10.30 | 100.00 |

Pabak index (Kappa adjusted for prevalence bias) = 0.84

## 4. Discussion

Results of this study show a linear positive association (P<0.001) between the score obtained with the Nova-UPF screener for Senegal and the dietary share of UPF obtained from the full 24-hour dietary recall, as the reference method There was near-perfect agreement between the score quintiles distribution of individuals and the UPF energy contribution quintiles.

The average percentage of calories provided by UPF (17.4%) was similar to that reported in France (17.4%) [27] and in Brazilian adults (17.7%) [28], and to other studies in Colombia (15.9%) [29] and Spain (17.3%) [30]. This reflects that consumption of these products occurs regardless of differences in dietary habits and economic development between countries. However, UPF consumption in the Senegalese context is still much lower than the rates reported in the USA [31], Canada [32], and Great Britain [8], where the availability of these products in the food supply is much higher [10–33].

The most consumed UPF category was “Bouillons, dipping sauce, vinaigrette or industrial salad dressings”. This is due to the presence of bouillons in this category, which is widely consumed by the majority (90%) of the Senegalese households’ [34], to enhance the taste of prepared dishes. The same happens for the category of salty sauces (Industrial mayonnaise, ketchup or mustard). The frequency of use of foods belonging to the categories “Instant milk powder or instant chocolate powder” and “Margarine” is mainly due to the habit of people to consume instant milk powder with coffee in the morning and bread coated with margarine, for breakfast. The consumption of “Chocolate bars, chocolate candies, confectionery or chewy products” is mostly due to snacking behaviour between meals, especially among younger people.

The absence of consumption of ’Industrial ice cream or popsicles’ could be attributed to the data collection period, which took place in winter, whereas these products are generally more consumed during hot weather (summer). Despite their high availability on the market, it appeared that ’Fruit-flavoured drinks prepared from a powdered mixture’ and ’Industrial jams or marmalades’ were not consumed by our study sample. This may be due to the fact that we have not caught profiles of people who consume these products, or because their consumption is more common in areas other than those covered by our study (e.g., rural or peri-urban). Applying the Nova tool at national level will give us a clearer assessment on consumption of all categories, and possibly allow us to adjust the adapted content further on.

The scores obtained and their distribution were very similar to results from the validation tool in Brazil. Indeed, in both studies, the most common scores were 1, 2, 3, and 4 [15]. However, the average percentage of energy contribution according to the score quintiles was lower for Senegal. It varied between 2.8% for the first quintile and 30.4% for the fifth quintile, while the Brazilian average contribution varied between 19.3% and 43.9%, respectively [15]. When considering the distribution of individuals according to Nova score quintiles and quintiles of energy contribution of UPF, the agreement between the Nova-UPF screener for Senegal and the 24h recall is higher in the first three quintiles, while the original Brazilian tool highlights more the extreme quintiles (first and fifth). These differences could be explained by the difference in dietary patterns and food culture between Senegal and Brazil, and the sample characteristics, such as age, sex, and education level, which can influence the consumption of UPF in quantity and variety.

Beyond the significant association, the concordance expressed through the Pabak index is better for the adapted version in Senegal compared to the original one [15]. Adaptation from a predefined original content allows for a more precise and exhaustive adjustment of the tool’s content. This suggests good results when considering that the Nova-UPF screener can be adapted in other countries, or its content can be updated in the future, according to changes in dietary habits.

The sample size of this study is unrepresentative, but this has no impact on the results because this study compares two methods to estimate UPF consumption in the same population. It was conducted based on probability sampling with a balanced sex-ratio population whose socio- demographic characteristics (level of education, age, profession, etc.) cover the entire concerned population. This is the first study on food consumption in Senegal using the Nova food classification and gives a relatively good idea of what the situation could be at the national level.

## 5. Conclusions

The Nova-UPF screener for Senegal is valid for measuring and monitoring the consumption of ultra-processed foods at the population level and over time. The score obtained with this tool accurately reflects the level of energy intake from these targeted foods. However, it will be necessary to perform this tool on a larger sample to better appreciate its applicability in nutritional surveillance and monitoring system.

## Data Availability

All data produced in the present study are available upon reasonable request to the authors

